# Validity and Limitations of the Empatica E4 Wristband for Autonomic and Thermoregulatory Sleep Monitoring Against Concurrent Polysomnography: A Wearanize+ Dataset Study

**DOI:** 10.64898/2026.06.10.26355348

**Authors:** Yildiz Dilara Parry, Giovanni Briganti

## Abstract

The Empatica E4 wristband provides continuous multi-modal physiological monitoring including blood volume pulse (BVP), electrodermal activity (EDA) and skin temperature (TEMP) but its validity for sleep-stage-specific autonomic and thermoregulatory monitoring has not been systematically evaluated against concurrent polysomnography (PSG). Using the Wearanize+ dataset which provides synchronised PSG, Empatica E4, and Zmax EEG recordings from 100 home-recorded participants; a systematic validation of Empatica E4 physiological signals against PSG ground truth across five sleep stages was conducted. Of 100 participants, 92 had Empatica data; 69 met Zmax EEG signal quality criteria and formed the analysis sample. Heart rate (HR) from the pre-computed Empatica HR channel showed valid stage-specific patterns (Wake: 70.9 bpm, N3: 61.2 bpm) and moderate inter-device MeanNN correspondence with PSG ECG (Spearman r=0.35-0.42 across stages). Skin temperature showed the expected thermoregulatory pattern (Wake: 33.92°C, N3: 35.48°C) and is recommended for downstream analyses. Tonic EDA showed an inverted stage pattern attributable to wrist sweat accumulation during deep sleep, representing a known confound for wrist-worn EDA during sleep. Phasic EDA showed plausible patterns and may be used with caution. These findings establish a validated feature set for Empatica E4 sleep research and directly inform multimodal psychiatric biomarker studies using the Wearanize+ dataset.

## 1. Introduction

Wearable physiological monitoring during sleep offers the potential to capture autonomic, thermoregulatory, and motor signatures of sleep health at population scale (Chinoy et al., 2021). The Empatica E4 wristband has become one of the most widely used research-grade wearable devices, providing blood volume pulse (BVP) at 64 Hz, electrodermal activity (EDA) at 4 Hz, skin temperature (TEMP) at 4 Hz and pre-computed heart rate (HR) at 1 Hz from a single wrist-worn sensor. These signals, if validated for sleep-stage-specific use, would provide access to autonomic (HRV), sympathetic arousal (EDA), accelerometry (ACC) and thermoregulatory (TEMP) dimensions of sleep physiology without the constraints of laboratory polysomnography (Föll et al., 2021; Mishra et al., 2018, 2020).

On the other hand, the physiological signals targeted by wrist-worn devices carry substantial clinical and scientific relevance for sleep research. Heart rate variability reflects the balance between sympathetic and parasympathetic autonomic regulation across sleep stages, with rmssd peaking during slow-wave sleep and declining during rem as sympathetic tone partially recovers (Burgess et al., 1997; Tobaldini et al., 2013). Reduced nocturnal HRV is associated with cardiovascular risk, insomnia disorder, and major depressive disorder, making it a candidate biomarker for sleep-related psychiatric and somatic vulnerability (Tobaldini et al., 2013). Skin temperature reflects peripheral thermoregulatory control mediated by the sympathetic nervous system: as core body temperature drops at sleep onset, peripheral vasodilation increases distal skin temperature, a mechanism whose disruption has been linked to insomnia and mood disorders (Harding et al., 2019; Kräuchi et al., 2000). Electrodermal activity captures sympathetic skin conductance responses that index autonomic arousal and emotional processing during sleep, with phasic EDA peaks occurring most frequently during REM and slow-wave sleep (Sano et al., 2014).

Wrist accelerometry provides a continuous measure of nocturnal movement, with progressive suppression from wakefulness through NREM sleep reflecting motor quiescence during deep sleep which is a pattern useful for gross sleep-wake discrimination and epoch quality assessment in downstream analyses (van Hees et al., 2013).

If these signals can be reliably captured by a wrist-worn device during unattended home sleep recordings, they would substantially expand the scale and ecological validity of autonomic sleep research and increase accessibility of sleep studies, downstreaming to mainstream smartwatches.

Prior validation studies of the Empatica E4 have focused predominantly on waking physiological tasks such as stress induction, emotion elicitation, cognitive load where signal quality is typically higher than during unattended overnight sleep (Aaslestad et al., 2026; Lee et al., 2026; Meegahapola et al., 2026; Shah et al., 2026). Wrist-worn PPG during sleep presents specific challenges not present in waking use: postural changes reduce perfusion at the wrist, the arm position during sleep varies unpredictably, and thermoregulatory sweating during deep sleep confounds EDA measurements (Zhang et al., 2019). Accelerometry during sleep presents its own interpretive challenge: the raw Euclidean norm of the three-axis signal is dominated by gravitational acceleration, meaning epoch-level standard deviation rather than mean magnitude is the appropriate movement feature for sleep characterisation (van Hees et al., 2013). The extent to which these factors compromise Empatica E4 validity for sleep monitoring has not been systematically characterised in a large concurrent PSG dataset.

The present study addresses five specific validation questions: (1) does Empatica pre-computed HR provide valid stage-specific cardiac estimates relative to PSG ECG; (2) are BVP-derived HRV metrics valid for sleep-stage-specific analyses; (3) is wrist EDA a valid index of autonomic arousal during sleep; (4) does wrist skin temperature provide reliable thermoregulatory characterisation across sleep stages; (5) does wrist accelerometry capture the expected motor quiescence pattern across sleep stages; and (6) are the feature-validity conclusions robust when PSG manual stage labels are replaced by wearable-derived automated staging, enabling a fully PSG-free deployment?

The Wearanize+ dataset provides the empirical conditions for this validation: synchronised full PSG and Empatica E4 recordings from 92 home-recorded participants, with PSG ECG providing gold-standard cardiac ground truth and PSG manual stage labels providing epoch-level staging accuracy. The present study, the third in a validation series using the Wearanize+ PlugNPlay dataset, evaluates Empatica E4 HR, HRV, EDA, TEMP and ACC against concurrent PSG reference measures across five sleep stages, and establishes a validated feature set for downstream multimodal sleep biomarker research.

## 2. Methods

### 2.1 Dataset and Participants

The Wearanize+ PlugNPlay dataset (Sikder et al., 2026) was used for this study. In total, 130 healthy participants aged 18-39 years (mean=23.16 years, SD=4.34; 89 female, 116 right-handed) were recruited via the Radboud University SONA participation system and completed one night of home recording. Inclusion criteria applied by the original dataset creators included: age 18-50 years, BMI ≥17.5, absence of known sleep disorder or mental illness, no sleep-affecting medication, no history of head or brain surgery, no epilepsy diagnosis, and not pregnant. The study was conducted under Donders Centre for Cognitive Neuroimaging blanket approval (NL45659.091.14, METC Oost-Nederland 2014/288) and in accordance with the Declaration of Helsinki. All recordings were performed in participants’ own homes without laboratory supervision.The PlugNPlay v1.0 release provides a processed, synchronised, and temporally truncated version of the raw recordings consolidated into a single EDF file per participant. All device signals are stored with Float32 precision, labelled according to the convention [device_ID]_[channel_name], and aligned to a common time grid. PSG-based manual and automated sleep scores are integrated into each EDF file as continuous channels at 1/30 Hz, corresponding to one score per 30-second epoch. The release includes 100 of the original 130 participants for whom both PSG and Zmax data were available and manual sleep scoring could be completed. PSG was recorded using SOMNOscreen Plus or Mentalab Explore Pro systems at 256 Hz, with ECG recorded via standard patch electrode placement. Empatica and PSG recordings were manually synchronised using accelerometer-based visual inspection as described in Sikder et al. (2026). Signals were extracted from the EDF files using Python’s MNE library and converted to numpy arrays for downstream processing.

Of the 100 PlugNPlay participants, 92 had Empatica E4 data available. Signal quality screening of concurrent Zmax EEG recordings (Parry & Briganti, 2026a) identified 74 participants with adequate EEG quality (OK group), of whom 69 had concurrent Empatica E4 data and formed the primary analysis sample.

Two additional subjects were excluded from HRV analyses due to absent PSG ECG channel, yielding N=67 for cardiac validation. All epoch selection was based exclusively on the PSG manual score channel, consistent with the recommendations of the companion staging and spectral validation studies (Parry & Briganti, 2026a, 2026b).

### 2.2 Empatica E4 Signals

The Empatica E4 wristband includes five physiological continuous signals from the non-dominant wrist: blood volume pulse (BVP, photoplethysmography, 64 Hz), electrodermal activity (EDA, 4 Hz), skin temperature (TEMP, 4 Hz), tri-axial accelerometry (ACC, 32 Hz), and pre-computed heart rate (HR, 1 Hz). The HR channel is derived from BVP by the device firmware using proprietary peak detection and smoothing algorithms, and is distinct from post-hoc BVP peak detection applied in the present study. In the Wearanize+ PlugNPlay dataset, all Empatica signals are pre-synchronised with PSG recordings using accelerometer-based manual synchronisation as described in Sikder et al. (2026), and stored as continuous time series aligned to the same 30-second epoch grid as PSG manual sleep scores. Synchronisation quality was verified by confirming matched recording durations across devices (mean 8.98±0.23 hours). Signals were extracted from the EDF files using Python’s MNE library and converted to numpy arrays prior to feature extraction. All analyses used PSG-defined epoch boundaries exclusively, ensuring that stage-specific feature extraction was not influenced by Empatica-derived staging or movement detection.

### 2.3 PSG Reference Signals

PSG ECG (channel ECG 2, 256 Hz, recorded via standard patch electrodes) served as the cardiac ground truth for HRV validation. R-peaks were detected using NeuroKit2 ecg_process(Makowski et al., 2021), which applies bandpass filtering, Pan-Tompkins peak detection, and signal quality assessment before returning a cleaned R-peak index. Time-domain HRV metrics were computed from the resulting R-peak series using NeuroKit2 hrv_time, and frequency-domain metrics using hrv_frequency, following the standards of the Task Force of the European Society of Cardiology (Electrophysiology, 1996) and the tutorial recommendations (Pham et al., 2021). PSG manual stage labels derived from full overnight scoring by an experienced sleep scorer following AASM criteria were used for all epoch selection throughout the study, consistent with the methodological recommendations established in the companion staging and spectral validation papers(Parry & Briganti, 2026b, 2026a). Automated U-Sleep scores available in the PlugNPlay release were not used for epoch selection, as manual labels provide superior accuracy for stage-specific physiological feature extraction.

### 2.4 HRV Validation

BVP PPG peaks were detected using NeuroKit2 ppg_process (Makowski et al., 2021; Pham et al., 2021). For each PSG-defined sleep stage, time-domain HRV metrics were computed using NeuroKit2 hrv_time: RMSSD, MeanNN, and SDNN. Frequency-domain metrics (LF, HF, LF/HF ratio) were computed using NeuroKit2 hrv_frequency. Corresponding metrics were computed from PSG ECG R-peaks for the same stage epochs. Pearson and Spearman correlations between Empatica BVP-derived and PSG ECG-derived HRV were computed at the subject level per stage. Bland-Altman analysis quantified systematic bias and limits of agreement for RMSSD (Martin Bland & Altman, 1986). Physiological plausibility was assessed by examining the stage-specific pattern of HR and RMSSD (expected: HR lowest in N3; RMSSD highest in N3 (Tobaldini et al., 2013)).

### 2.5 EDA, Temperature, and Accelerometry

EDA signals were decomposed into tonic and phasic components using NeuroKit2 (Makowski et al., 2021) eda_process. Mean tonic EDA, mean phasic EDA, and phasic peak count were computed per 30-second epoch and averaged per stage per subject. Physiological validity was assessed by examining the stage-specific tonic EDA pattern (expected: lowest in N3 reflecting minimal sympathetic arousal during deep sleep) (Sano et al., 2014; Tobaldini et al., 2013).

Skin temperature means and standard deviations were computed per 30-second epoch and averaged per stage per subject. Physiological validity was assessed by examining the stage-specific pattern of mean wrist skin temperature, with expected progressive increase from Wake to N3 reflecting peripheral vasodilation as core body temperature drops at sleep onset as a well-established autonomic thermoregulatory mechanism by which heat is redistributed from core to distal skin surfaces during sleep (Harding et al., 2019; Kräuchi et al., 2000). Stage-specific means were confirmed against published normal ranges for wrist skin temperature during sleep (Martinez-Nicolas et al., 2013; Sarabia et al., 2008). Repeated-measures ANOVA with Greenhouse-Geisser correction was applied to test the significance of the stage-dependent temperature pattern, with Bonferroni-corrected post-hoc pairwise comparisons and bootstrap 95% confidence intervals computed for each stage mean. Epoch-level standard deviation of skin temperature was also extracted per stage as a measure of thermoregulatory instability, given its previously demonstrated relevance as an objective wearable biomarker of psychiatric symptom burden (Mason et al., 2024; Shin et al., 2025).

Wrist accelerometry magnitude from the Empatica E4 (32 Hz) was computed per 30-second epoch as the Euclidean norm of the three-axis signal Epoch-level standard deviation of accelerometry magnitude was used as the primary movement feature, as the raw Euclidean norm mean is dominated by gravitational acceleration and does not distinguish movement across stages at the mean level (Van Hees et al., 2015). Mean magnitude, standard deviation, and proportion of epochs exceeding a movement threshold were computed per stage per subject and averaged within each stage. Physiological validity was assessed by examining the stage-specific pattern of accelerometry standard deviation, with expected highest values during Wake and lowest during N3 reflecting motor quiescence during deep sleep.

### 2.6 Secondary validation under wearable-derived staging

To assess whether the feature-validity conclusions depend on PSG-defined sleep staging, all primary analyses were repeated using wearable-derived stage labels as a secondary validation. Automated sleep staging was performed on the Zmax EEG left channel (F7-Fpz) using YASA v0.6.5 (Vallat & Walker, 2021) applying the same per-subject MNE RawArray pipeline and N2-referenced calibration established in previous preprint (Parry & Briganti, 2026b). Preliminary findings on YASA automated staging achieves substantial agreement with PSG manual scoring in adequate-quality home recordings from this dataset as Cohen’s κ=0.450 (Parry & Briganti, 2026a), providing a validated PSG-free staging solution. Stage-specific Empatica E4 HR and skin temperature means were computed using YASA-defined epoch boundaries following the same extraction procedure described in Sections 2.4 and 2.5. The concordance between PSG-staged and YASA-staged feature estimates was assessed at the subject level using Spearman correlation.

## 3. Results

### 3.1 Heart Rate: Valid Stage Pattern

The Empatica pre-computed HR channel showed the expected monotonic decrease from Wake to N3: Wake=70.9 bpm, N1=66.0 bpm, N2=61.4 bpm, N3=61.2 bpm, REM=62.1 bpm. This pattern is consistent with progressive parasympathetic dominance from wakefulness through NREM sleep and slight sympathetic reactivation during REM (Burgess et al., 1997; Tobaldini et al., 2013). MeanNN derived from HR (60,000/HR in ms) showed the corresponding increase: Wake=872.8 ms to N3=1012.0 ms (Figure 1B). Heart rate showed a significant stage-dependent pattern (RM-ANOVA: F(3,195)=85.67, p<0.0001, η^2^=0.188; Greenhouse-Geisser correction applied due to sphericity violation, W=0.282, p<0.0001).

**Figure 1.**
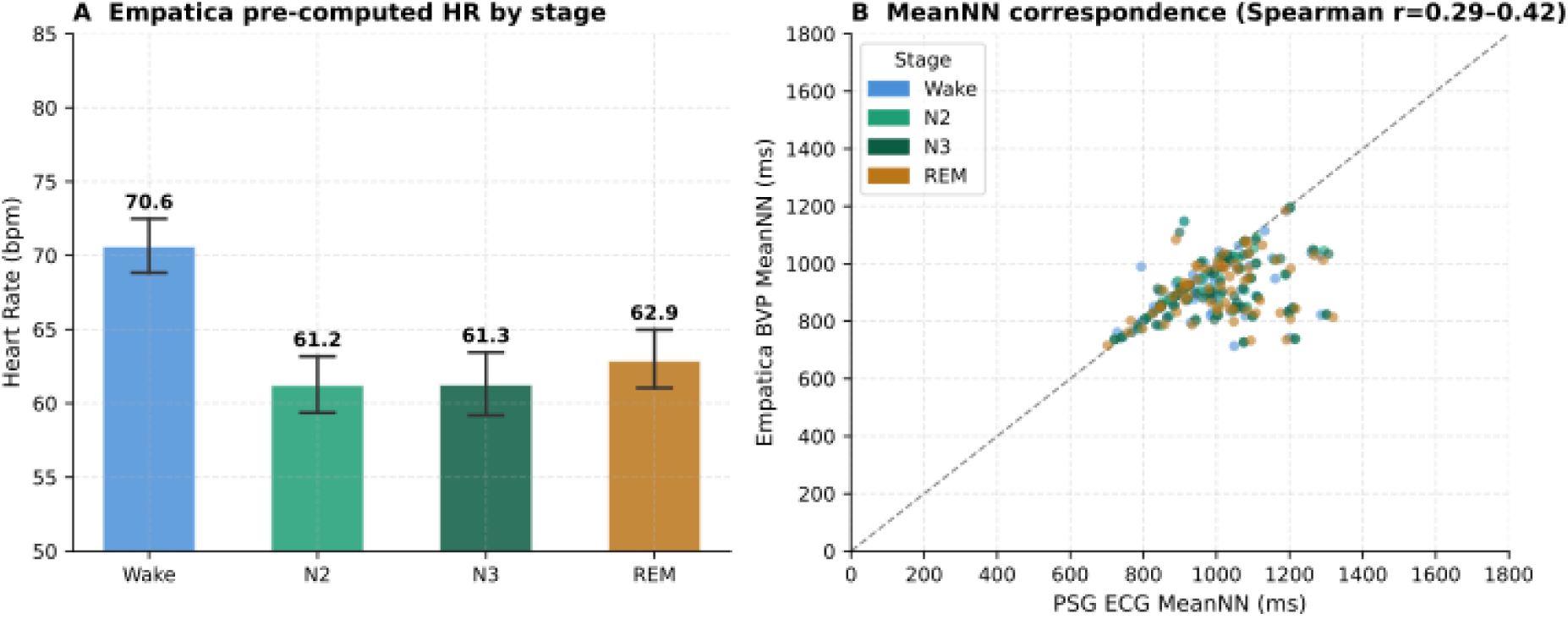
A & B. Heart rate validation: Empatica E4 vs PSG ECG

**Figure 2.**
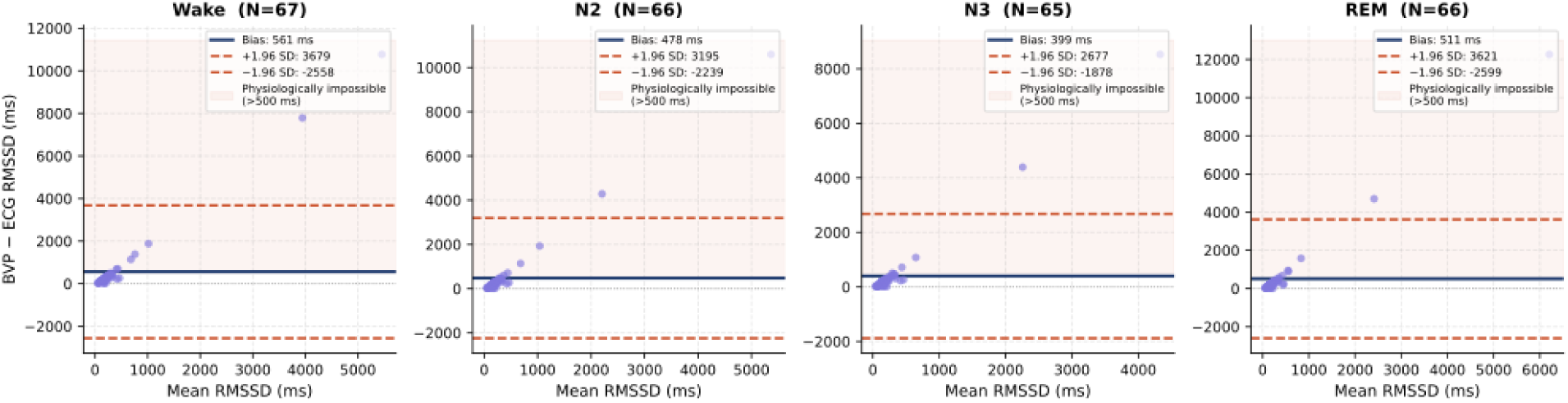
Bland-Altman analysis of RMSSD: Empatica BVP-derived versus PSG ECG-derived across four sleep stages.

Bootstrap 95% confidence intervals confirmed stage-specific HR estimates: Wake 70.6 bpm [68.8-72.4], N2 61.2 bpm [59.3-63.2], N3 61.3 bpm [59.2-63.5], REM 62.9 bpm [61.0-65.0] as shown on Figure 1A. Bonferroni-corrected post-hoc comparisons revealed that Wake HR was significantly higher than all sleep stages (all p<0.0001, Hedges g=0.94–1.21), while N2 and N3 did not differ significantly (p=1.000, g=0.011), and REM was intermediate between Wake and NREM stages (REM vs Wake: p<0.0001, g=0.94; REM vs N2: p<0.001, g=0.23). This stage-specific pattern is consistent with parasympathetic predominance during NREM and partial autonomic reactivation during REM sleep.

These values are physiologically normal and indicate that the Empatica HR channel provides valid stage-specific heart rate estimates.

### 3.2 BVP-Derived HRV: Not Valid Due to Wrist PPG Artefacts

BVP peak detection produced physiologically impossible HRV values across all stages and all subjects. Bland-Altman analysis of RMSSD revealed systematic positive bias (BVP minus ECG): Wake=+560.9 ms, N2=+477.6 ms, N3=+399.3 ms, REM=+511.1 ms. These biases represent RMSSD overestimates of 400–560 ms; values that are physiologically impossible (normal sleep RMSSD is 20-100 ms) (Electrophysiology, 1996; Tobaldini et al., 2013). The limits of agreement were extremely wide (-2,557 to +3,679 ms for Wake), indicating near-complete absence of inter-device agreement. The stage pattern of BVP-derived RMSSD was inverted relative to ECG (Wake highest, N3 lowest), the opposite of the expected physiological pattern.

After applying physiological plausibility filters (RMSSD <200 ms; MeanNN 400-2000 ms), 45.1% of epochs survived. Even in this cleaned subset, the stage pattern remained inverted (Wake=104.6 ms, N3=86.0 ms), confirming that the problem is not limited to extreme outliers but reflects systematic BVP peak detection failure during sleep. The cause is wrist PPG motion artefact: during sleep, postural changes, thermoregulatory skin perfusion changes, and periodic arousals produce large-amplitude BVP oscillations that the peak detector misidentifies as cardiac pulses, generating spurious short inter-beat intervals (Charlton et al., 2025; Van Gent et al., 2019; Zhang et al., 2019).

MeanNN showed moderate Spearman correlations with PSG ECG MeanNN (Wake: r=0.351, p=0.004; N2: r=0.380, p=0.002; N3: r=0.421, p=0.001; REM: r=0.289, p=0.018), indicating that rank-order correspondence is partially preserved despite absolute value invalidity. Pearson correlations were near-zero or negative for all metrics except MeanNN, driven by outlier contamination.

**Table 1.**
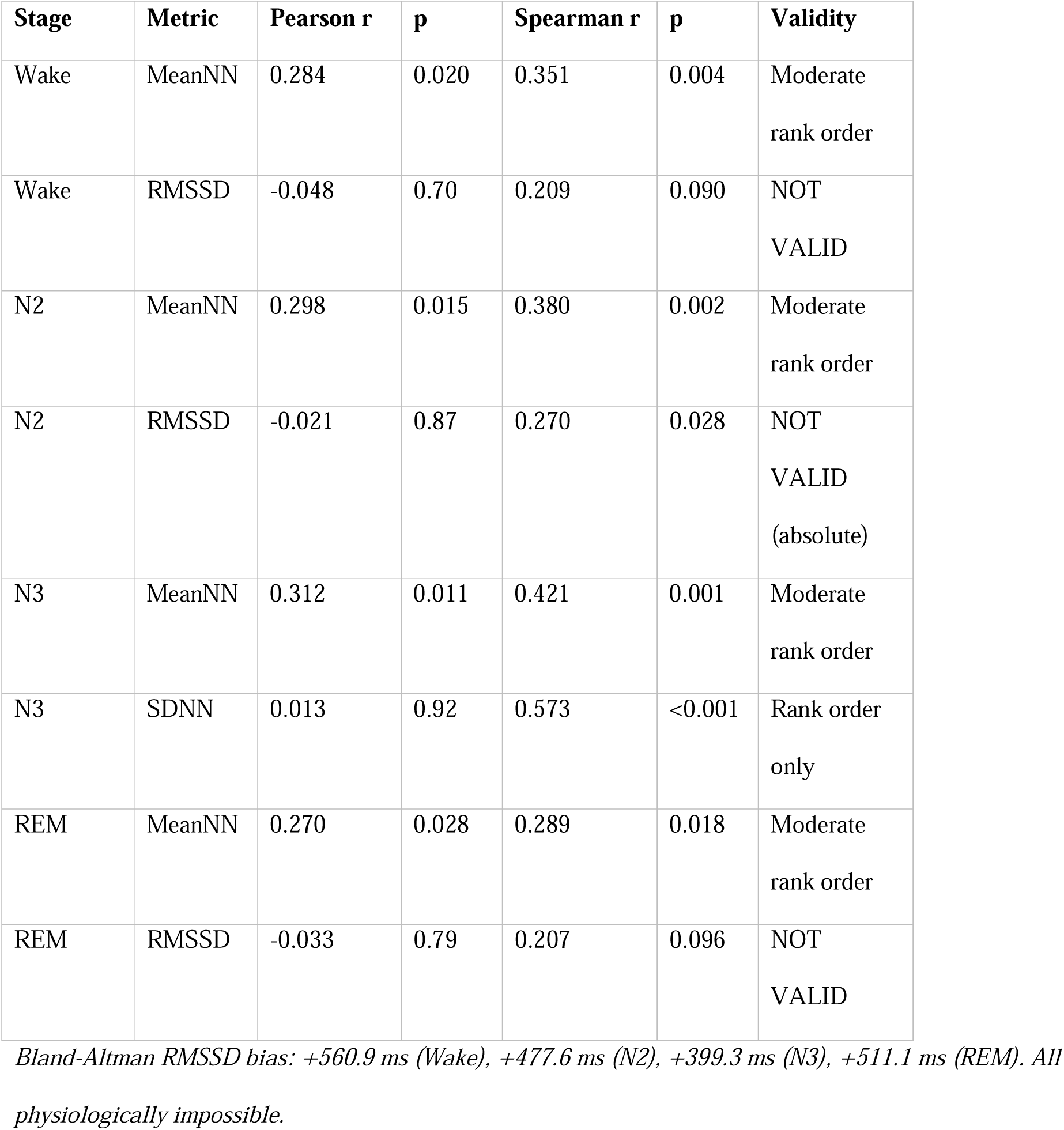
HRV validation: Empatica BVP vs PSG ECG (N=67 subjects).

| Stage | Metric | Pearson r | p | Spearman r | p | Validity |
| --- | --- | --- | --- | --- | --- | --- |
| Wake | MeanNN | 0.284 | 0.020 | 0.351 | 0.004 | Moderate<br>rank order |
| Wake | RMSSD | -0.048 | 0.70 | 0.209 | 0.090 | NOT<br>VALID |
| N2 | MeanNN | 0.298 | 0.015 | 0.380 | 0.002 | Moderate<br>rank order |
| N2 | RMSSD | -0.021 | 0.87 | 0.270 | 0.028 | NOT<br>VALID<br>(absolute) |
| N3 | MeanNN | 0.312 | 0.011 | 0.421 | 0.001 | Moderate<br>rank order |
| N3 | SDNN | 0.013 | 0.92 | 0.573 | <0.001 | Rank order<br>only |
| REM | MeanNN | 0.270 | 0.028 | 0.289 | 0.018 | Moderate<br>rank order |
| REM | RMSSD | -0.033 | 0.79 | 0.207 | 0.096 | NOT<br>VALID |

*Bland-Altman RMSSD bias: +560.9 ms (Wake), +477.6 ms (N2), +399.3 ms (N3), +511.1 ms (REM). All physiologically impossible.*

### 3.3 Skin Temperature: Valid

Skin temperature showed the expected thermoregulatory pattern across all stages: Wake= 33.92°C (±4.02), N1=35.06°C (±1.30), N2=35.40°C (±1.03), N3=35.48°C (±1.00), REM=35.43°C (±0.83). The stage-dependent pattern was statistically significant (RM-ANOVA: F(3,195)=12.42, p<0.0001, η^2^=0.124; Greenhouse-Geisser correction applied due to sphericity violation, W= 0.001, corrected p= 0.001). Figure 3A visualises the confirmed Bootstrap 95% confidence intervals stage-specific estimates: Wake 33.79°C [32.80-34.41], N2 35.39°C [35.27-35.51], N3 35.44°C [35.26-35.61], REM 35.44°C [35.28-35.59].

**Figure 3A and 3B.**
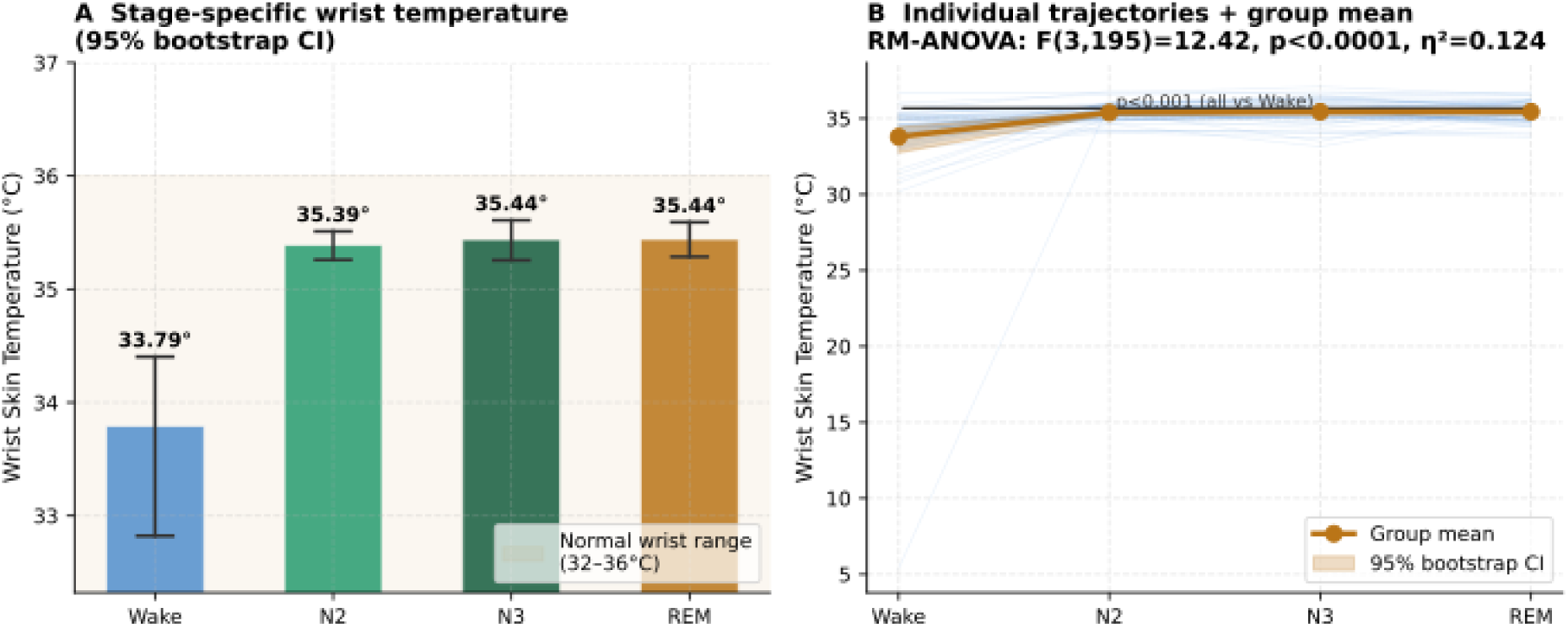
Wrist skin temperature validation as stage-specific pattern (N=69)

Bonferroni-corrected post-hoc comparisons showed that Wake temperature was significantly lower than all sleep stages (Wake vs N2: p=0.005, g=0.61; Wake vs N3: p=0.004, g=0.63; Wake vs REM: p=0.004, g=0.63), while N2, N3, and REM did not differ significantly from each other (all p=1.000) (Figure 3B). Temperature rises progressively from Wake to N3, consistent with peripheral vasodilation as core body temperature drops at sleep onset (Harding et al., 2019; Kräuchi et al., 2000). The larger Wake SD (4.02°C) reflects within-subject variability during nocturnal arousals and pre-sleep wakefulness. Temperature values are physiologically normal; 32–36°C at the wrist (Martinez-Nicolas et al., 2013; Sarabia et al., 2008). Skin temperature is validated for downstream sleep stage-specific analyses and is recommended as a primary Empatica feature for thermoregulatory research.

### 3.4 EDA: Tonic Confounded, Phasic Usable with Caution

Tonic EDA showed an inverted stage pattern: Wake=1.13 μS (±2.09), N1= 1.38 μS (±2.64), N2=1.60 μS (±2.70), N3= 2.93 μS (±4.66), REM= 1.36 μS (±1.99). As shown on Figure 4A, tonic EDA was highest during N3 rather than lowest; the opposite of the expected pattern (Sano et al., 2014). This is a known confound of wrist-worn EDA during sleep: sweat accumulates under the wristband during periods of reduced movement and physical activity (particularly deep sleep), increasing apparent skin conductance through a hydration mechanism rather than sympathetic arousal (Milstein & Gordon, 2020; Van Der Mee et al., 2021). Tonic EDA during sleep therefore reflects skin hydration rather than autonomic sympathetic activity (Boucsein, 2014) and should not be used as a sympathetic arousal proxy in sleep studies.

**Figure 4A and 4B.**
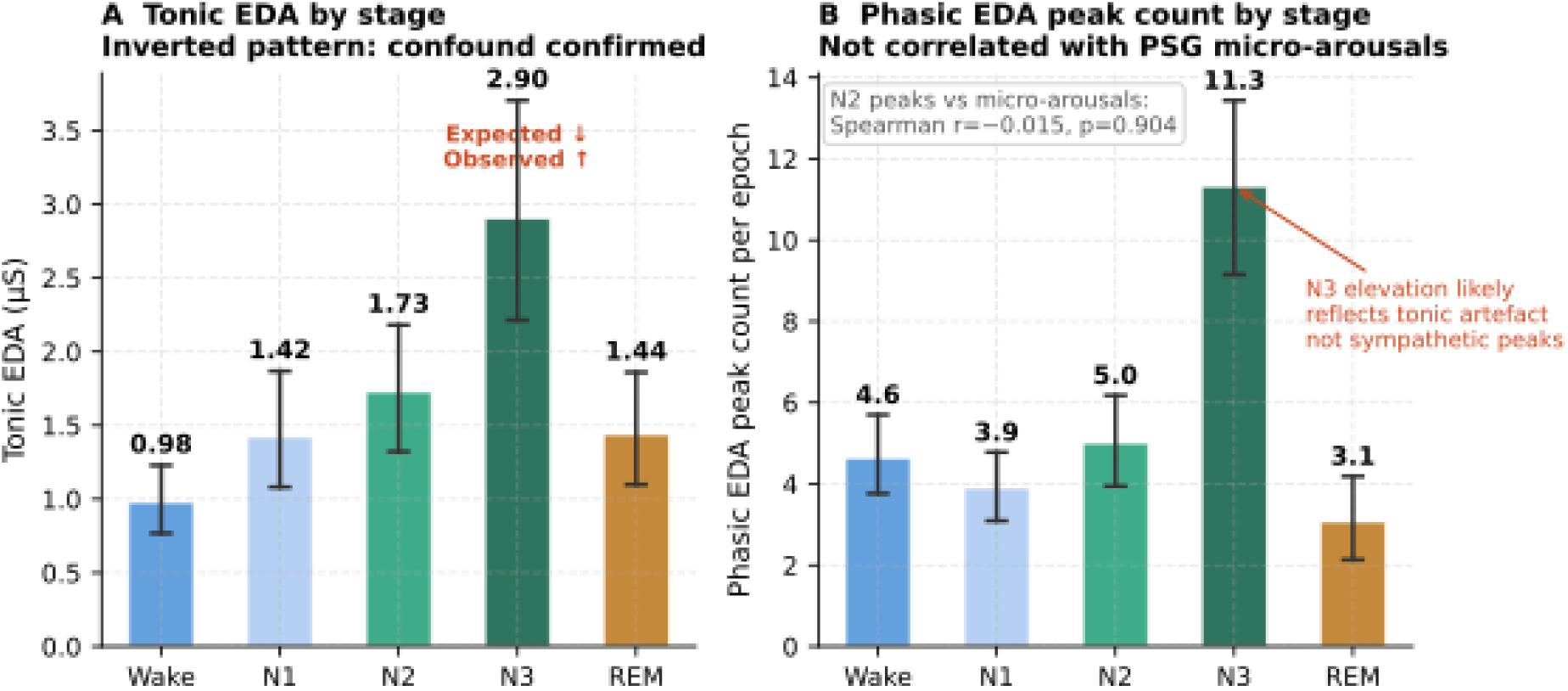
Comparison of tonic confound vs phasic features of electrodermal activity validation (N=69)

Phasic EDA reflects discrete sympathetic skin responses superimposed on the tonic baseline (Figure 4B). While no PSG equivalent exists for direct validation of phasic EDA, internal consistency was assessed by correlating phasic EDA peak count during N2 with the number of micro-arousals, defined as single Wake epochs flanked by NREM epochs as a proxy for cortical arousal fragmentation. No significant association was found (Spearman r= -0.015, p= 0.904, N= 69). The absolute values and stage distribution of phasic EDA were physiologically plausible, and phasic EDA features may be used in downstream analyses with explicit acknowledgement that they have not been validated against a direct physiological reference standard and do not index micro-arousal frequency.

### 3.5 Wrist Accelerometry: Valid Movement Pattern

Wrist accelerometry epoch-level standard deviation showed the expected stage-dependent pattern: Wake=2.23±0.90, N2=0.38±0.09, N3=0.33±0.09, REM=0.46±0.17. Movement variability was highest during Wake and lowest during N3, consistent with progressive motor quiescence from wakefulness through NREM sleep, with a slight increase during REM reflecting phasic motor activity characteristic of that stage.

Mean accelerometry magnitude did not differ meaningfully across stages (range 63.93 to 64.61 arbitrary units), confirming that epoch standard deviation rather than mean magnitude is the appropriate feature for movement characterisation in this dataset. The elevated Wake accelerometry standard deviation was independent of Wake epoch count across subjects (Spearman r=-0.157, p=0.198, N=69), confirming that the wake movement pattern reflects genuine physiological wakefulness rather than a recording duration artefact. Wrist accelerometry standard deviation is therefore suitable for movement-based epoch quality screening and gross sleep-wake discrimination in downstream analyses, and is included in the validated feature set with this application in mind.

### 3.5 Feature validation under wearable-derived staging

Heart rate and skin temperature patterns were preserved under YASA automated staging. HR showed the expected stage-dependent pattern: Wake=65.29 bpm, N2=61.21 bpm, N3=61.97 bpm, REM=62.69 bpm, with Wake elevated above all sleep stages consistent with the PSG-staged result (PSG: Wake=69.69, N2=61.24, N3=61.27, REM=62.87 bpm). Mean differences between PSG-staged and YASA-staged HR estimates were negligible during sleep stages (N2: -0.04 bpm, N3: +0.69 bpm, REM: -0.19 bpm). The larger Wake difference (-4.40 bpm) reflects the known tendency of automated staging to classify fewer pre-sleep and post-sleep wakefulness epochs as Wake, resulting in YASA Wake epochs being weighted toward quiescent nocturnal arousals with lower HR than extended wakefulness periods captured by PSG (Schyvens et al., 2025). Subject-level agreement between PSG-staged and YASA-staged HR was excellent across all stages (Wake: r=0.861, N2: r=0.986, N3: r=0.954, REM: r=0.973; all p<0.0001, N=66–69).

Skin temperature retained the expected peripheral vasodilation pattern under automated staging (Wake=34.51°C, N2=35.41°C, N3=35.49°C, REM=35.41°C), with mean differences from PSG-staged estimates of 0.02-0.71°C across stages. As with HR, the Wake difference (+0.71°C) reflects YASA’s tendency to include fewer cold pre-sleep epochs in the Wake category, shifting the YASA Wake temperature estimate slightly upward. Subject-level agreement between PSG-staged and YASA-staged temperature was high across all stages (Wake: r=0.762, N2: r=0.951, N3: r=0.956, REM: r=0.888; all p<0.0001, N=66-69).

Skin temperature standard deviation showed robust concordance under automated staging. Mean differences between PSG-staged and YASA-staged TEMP std estimates were negligible across all sleep stages (N2: - 0.0002, N3: +0.0002, REM: +0.0004°C), with a slightly larger Wake difference (-0.0099°C) consistent with the wakefulness epoch classification effect described above. Subject-level agreement was high across all stages (Wake: r=0.813, N2: r=0.885, N3: r=0.826, REM: r=0.855; all p<0.0001, N=66-69). Phasic EDA peak counts showed preserved stage-level patterns under automated staging. Mean differences between PSG-staged and YASA-staged phasic peak estimates were small across sleep stages (N2: -0.22, N3: -0.08, REM: +0.25 peaks per epoch), with a slightly larger Wake difference (+0.50 peaks per epoch) consistent with the wakefulness epoch classification effect. Subject-level concordance was high during sleep stages (N2: r=0.946, N3: r=0.879, REM: r=0.899; all p<0.0001) but moderate during Wake (r=0.538, p<0.0001), indicating that phasic EDA counts during sleep stages are robust to the source of stage labels, while Wake estimates are more sensitive to staging method.Wrist accelerometry standard deviation retained the expected movement suppression pattern under automated staging (Wake=1.23, N2=0.36, N3=0.43, REM=0.44), with the correct direction where the Wake highest, sleep stages substantially lower was preserved under both staging methods. Mean differences during sleep stages were small (N2: -0.016, REM: -0.023), though the N3 difference was slightly larger (+0.094), likely reflecting YASA’s known tendency to over-call N3 relative to PSG manual scoring in this dataset (Parry & Briganti, 2026a) which may introduce some N2 epochs with marginally higher movement into the YASA N3 pool. Subject-level concordance was moderate across all stages (Wake: r=0.544, N2: r=0.704, N3: r=0.620, REM: r=0.599; all p<0.0001, N=66-69), lower than HR and TEMP but still consistently significant. The Wake ACC std difference (-1.001) again reflects the wakefulness epoch classification effect rather than a feature instability.

The feature-validity conclusions for all signal types: HR valid, skin temperature valid, BVP-derived HRV invalid, tonic EDA confounded, which were unchanged when PSG manual staging was replaced by YASA automated staging.

**Table 2.** Empatica E4 feature validity summary for sleep stage-specific analyses.

| Feature | Stage pattern<br>(PSG-staged) | Stage pattern<br>(YASA-staged) | PSG vs YASA<br>Concordance (r) | Validity | Recommendation |
| --- | --- | --- | --- | --- | --- |
| **HR (pre-computed, 1 Hz)** | <u>Correct</u><br>(Wake=69.7,<br>N2=61.2,<br>N3=61.3,<br>REM=62.9 bpm) | <u>Correct</u><br>(Wake=65.3,<br>N2=61.2,<br>N3=62.0,<br>REM=62.7 bpm) | Wake: 0.861;<br>N2: 0.986;<br>N3: 0.954;<br>REM: 0.973<br>(all p<0.0001) | Valid | Use directly; derive<br>MeanNN=60000/HR |
| **BVP → RMSSD** | <u>Inverted</u><br>(Wake>N3) | Not repeated: PSG<br>*conclusion*<br>*sufficient for*<br>*exclusion* | - | NOT<br>VALID | Do not use; wrist PPG<br>artefacts |
| **BVP → MeanNN** | Correct rank order | Not repeated | - | Rank<br>order only | Spearman correlations<br>only; not absolute |
| **BVP → LF/HF** | Not validated | Not repeated | - | NOT<br>VALID | Do not use |
| **TEMP mean** | <u>Correct</u><br>(Wake=33.79°C<br>rising to<br>N3=35.44°C) | <u>Correct</u><br>(Wake=34.51°C,<br>N2=35.41°C,<br>N3=35.49°C,<br>REM=35.41°C) | Wake: 0.762;<br>N2: 0.951;<br>N3: 0.956;<br>REM: 0.888<br>(all p<0.0001) | Valid | Use directly per stage |
| **TEMP std (variability)** | <u>Correct</u> (Wake<br>highest) | <u>Correct</u> (Wake<br>highest; differences<br><0.01 across sleep<br>stages) | Wake: 0.813;<br>N2: 0.885;<br>N3: 0.826;<br>REM: 0.855<br>(all p<0.0001) | Valid | Use per stage |
| **EDA tonic** | <u>Inverted</u> (N3<br>highest) | Not repeated: PSG<br>*conclusion*<br>*sufficient for*<br>*exclusion* | - | NOT<br>VALID | Wrist hydration<br>confound during deep<br>sleep |
| **EDA phasic** | <u>Correct</u><br>(Wake=7.26,<br>N2=8.49,<br>N3=8.02, | <u>Correct</u><br>(Wake=7.76,<br>N2=8.28,<br>N3=7.94, | Wake: 0.538;<br>N2: 0.946;<br>N3: 0.879;<br>REM: 0.899 | Not PSG<br>valid | Use with caution; sleep<br>stage phasic peaks<br>robust to staging<br>source; Wake estimate |
| | *REM=7.90 per epoch)* | *REM=8.14 per epoch)* | (all $p<0.0001$) | | less reliable |
| **ACC std (wrist)** | <u>Correct</u><br>(*Wake=2.23, N2=0.38, N3=0.33, REM=0.46*) | <u>Correct</u><br>(*Wake=1.23, N2=0.36, N3=0.43, REM=0.44*) | Wake: 0.544;<br>N2: 0.704;<br>N3: 0.620;<br>REM: 0.599<br>(all $p<0.0001$) | Valid for movement screening | Use epoch SD not mean magnitude;<br>Wake estimate sensitive to staging method |

## 4. Discussion

### 4.1 The Fpz reference problem and BVP artefacts: parallel limitations

The two primary validity failures identified in the Wearanize+ validation series share a common underlying cause: the geometry of sensor placement during sleep. The Fpz active reference electrode attenuating Zmax EEG power (Parry & Briganti, 2026b) and wrist PPG motion artefacts invalidating Empatica BVP-derived HRV both arise because the sensor is positioned where the biological signal is attenuated or contaminated by non-target sources. In the Zmax case, the active Fpz reference electrode sits over prefrontal cortex and partially cancels the EEG signal it is intended to measure. In the Empatica case, the wrist PPG sensor is positioned at a site where nocturnal postural changes, thermoregulatory perfusion shifts, and movement artefacts produce optical signal disturbances that the peak detector cannot distinguish from cardiac pulses (Zhang et al., 2019).

This parallel has practical implications for wearable device design and data analysis. The Fpz reference problem is correctable post-hoc through per-subject N2 calibration, on the other hand; the wrist PPG problem is not correctable post-hoc without a concurrent cardiac reference signal, because once false peaks have been inserted into the IBI series they cannot be reliably distinguished from true cardiac intervals. This asymmetry suggests a design hierarchy: EEG-based features from the Zmax can be salvaged through calibration, while BVP-derived HRV from the Empatica requires hardware-level solutions such as firmware-based artefact rejection using the proprietary IBI file, adaptive filtering algorithms, or signal quality indexing before peak detection (Van Gent et al., 2019).

Future wearable devices targeting sleep HRV monitoring should prioritise signal quality assessment as a first-pass filter before any HRV computation, and researchers should treat wrist PPG HRV as unvalidated in sleep contexts until device-specific validation against concurrent ECG is available.

### 4.2 Temperature as a valid autonomic proxy

Skin temperature provides a physiologically valid, artefact-robust measure of peripheral thermoregulation across sleep stages. The stage-specific pattern of 33.92°C at Wake rising to 35.48°C at N3 is consistent with the established thermoregulatory model of sleep: as core body temperature drops at sleep onset, peripheral vasodilation mediated by the sympathetic nervous system increases distal skin temperature (Harding et al., 2019; Kräuchi et al., 2000). This mechanism makes wrist temperature an indirect but valid proxy for autonomic thermoregulatory control during sleep, capturing the same peripheral vascular response that drives the well-documented association between distal skin warming and sleep onset latency.

Beyond its role as a thermoregulatory marker, wrist skin temperature has emerging relevance as a psychiatric biomarker with direct translational implications. Previous examinations reported associations between wearable sensor-assessed distal body temperature and self-reported depressive symptoms in over 20,000 participants over approximately seven months as part of the TemPredict Study, finding that greater depression symptom severity was associated with higher wearable-assessed body temperature and lower diurnal temperature amplitude (Mason et al., 2024). Also demonstrated previously; 24-hour wrist skin temperature rhythms are significantly blunted and less stable in young people with emerging mood disorders compared to healthy controls (Shin et al., 2025).

These converging findings position wrist skin temperature as the most immediately clinically translatable Empatica feature for psychiatric sleep research. Its validity established here, combined with its demonstrated association with depressive symptomatology across independent cohorts, supports its prioritisation as the primary wearable autonomic feature in clinical cohort extensions where autonomic thermoregulatory dysregulation is expected to be more pronounced than in the healthy community sample studied here.

### 4.3 EDA wrist confound during sleep

The inverted tonic EDA pattern during NREM sleep is a known and documented limitation of wrist-worn EDA during sleep (Sano et al., 2014) and subsequently reported in multiple Empatica validation studies (Milstein & Gordon, 2020; Van Der Mee et al., 2021). During N3, reduced movement and thermoregulatory sweating cause sweat to accumulate under the wristband without evaporating, increasing apparent skin conductance through a passive hydration mechanism rather than active sympathetic sweat gland stimulation (Kim et al., 2025; Van Der Mee et al., 2021). This confound is not correctable post-hoc without a concurrent sympathetic reference signal. Researchers using Empatica EDA for sleep studies should use phasic rather than tonic features, interpret EDA findings with explicit acknowledgement of this confound, and avoid using tonic EDA as a sympathetic arousal index during sleep.

Phasic EDA is less susceptible to this confound because it reflects rapid discrete sympathetic skin conductance responses rather than the slow-drifting tonic baseline that accumulates hydration artefact over minutes. However, as reported in Section 3.4, phasic EDA peak count did not correlate with PSG-defined micro-arousal frequency (r= -0.015, p= 0.904), indicating that phasic EDA does not index cortical arousal fragmentation and likely captures a distinct dimension of autonomic sleep physiology. Its use in downstream analyses is therefore conditional on explicit acknowledgement that it reflects an unvalidated autonomic signal of uncertain origin rather than a calibrated sympathetic arousal proxy.

### 4.4 Implications for multimodal sleep research

The validated Empatica feature set established here, HR-derived MeanNN, skin temperature, and phasic EDA, forms the physiological input layer for multimodal sleep biomarker analyses alongside the validated Zmax EEG spectral features (Parry & Briganti, 2026b). MeanNN Spearman correlations of 0.35-0.42 between Empatica HR-derived and PSG ECG-derived values indicate modest rank-order agreement that is sufficient for group-level analyses such as comparing mean MeanNN between diagnostic groups or correlating it with dimensional symptom scores but insufficient for individual-level clinical decisions, such as using a single subject’s wrist-derived MeanNN as a diagnostic threshold. Researchers should treat Empatica HR-derived cardiac features as population-level biomarkers and explicitly avoid their use for individual-level clinical inference until higher-resolution validation is available.

The systematic feature validity characterisation reported on this study has direct implications for the planned extension of this methodology to clinical psychiatric cohorts. In clinical populations with depression or anxiety disorders, autonomic dysregulation is expected to produce larger and more consistent stage-specific HR and temperature effects than observed in this healthy community sample, potentially enabling BVP-derived HRV metrics to achieve validity thresholds not reached here.

### 4.5 Robustness of feature validity with wearable-derived staging

The secondary validation addresses the fundamental question of whether the present framework requires PSG not only for initial signal quality screening but also for the staging that enables stage-specific interpretation. The results demonstrate that it does not. Feature-validity conclusions replicated fully when YASA automated staging replaced PSG manual labels, with subject-level HR and temperature concordance exceeding r=0.86 and r=0.76 respectively across all stages. The minor Wake discrepancy reflects a known property of automated staging, that brief nocturnal arousals are more reliably detected than extended pre-sleep wakefulness periods and does not affect the validity conclusions for sleep stages. These findings confirm that the Empatica E4 validation framework presented here is operable in a fully wearable deployment. Combined with the companion staging validation establishing YASA as a reliable automated staging solution for this dataset (Parry & Briganti, 2026a), the two papers together define a complete PSG-free sleep monitoring pipeline: automated staging provides the stage labels, and the present validation defines which physiological features can be trusted within each automatically-defined stage. A single concurrent PSG recording is required only for initial quality screening; all subsequent nights can be monitored using Zmax automated staging with validated Empatica E4 features.

### 4.6 Limitations

Several limitations should be noted. First, IBI files were not available in the Wearanize+ PlugNPlay parquet release, preventing evaluation of firmware-computed inter-beat intervals which may substantially outperform post-hoc BVP peak detection. The Empatica E4 firmware applies proprietary artefact rejection algorithms during IBI computation that are unavailable to post-hoc peak detectors operating on the raw BVP waveform. Researchers accessing raw Empatica data directly, rather than through pre-packaged datasets, should prioritise the IBI.csv file as the primary source for HRV computation rather than applying peak detection to the BVP channel. Firmware-derived IBI from the Empatica E4 shows substantially better agreement with ECG than post-hoc BVP peak detection in waking contexts (Schuurmans et al., 2020), suggesting that the invalidity reported here for BVP-derived RMSSD may not apply to firmware IBI in the same dataset. Future reanalysis of the Wearanize+ data using firmware IBI files obtained directly from raw Empatica exports is therefore a priority, and may recover meaningful HRV validity not achievable from the PlugNPlay BVP channel. Alternatively, adaptive PPG signal quality indices such as the SQI implemented in NeuroKit2 or HeartPy (Van Gent et al., 2019) can identify and exclude artefacted BVP segments before peak detection, potentially recovering valid HRV windows in a subset of subjects and epochs.Second, EDA phasic features were not validated against a PSG equivalent due to the absence of a gold-standard peripheral sympathetic reference in the PSG montage. The correlation between phasic EDA peaks and PSG-defined micro-arousals was null (r=0.015, p=0.904), indicating that phasic EDA does not index cortical arousal frequency, but its relationship to other autonomic sleep events such as autonomic arousals, REM phasic bursts, or K-complexes remains untested. Third, the healthy young adult cohort aged 18 to 39 years limits generalisability to populations with different autonomic profiles, including older adults in whom thermoregulatory capacity is reduced, clinical populations with autonomic dysregulation, and individuals with obesity in whom wrist PPG signal quality may differ systematically. Fourth, the single-night design precludes assessment of night-to-night reliability for any validated feature. Published evidence for EEG spectral features suggests moderate to good night-to-night reliability (Brunner et al., 1993) but equivalent data for wrist temperature and HR across nights in home recordings are not available and represent a priority for longitudinal validation studies.

## 5. Conclusions

This study provides the first systematic per-sleep-stage validation of the Empatica E4 wristband against concurrent PSG in a large home-recording dataset. Pre-computed HR and derived MeanNN are valid for stage-specific cardiac monitoring. Skin temperature provides physiologically valid thermoregulatory characterisation across sleep stages and is the most robust Empatica feature for downstream sleep biomarker research. BVP-derived beat-to-beat HRV metrics (RMSSD, SDNN, LF/HF) are not valid for absolute quantification due to systematic wrist PPG artefacts, though rank-order MeanNN correspondence is partially preserved. Tonic EDA is confounded by wrist sweat accumulation during deep sleep and should not be used as a sympathetic arousal proxy. These findings establish an evidence-based feature selection framework for Empatica E4 sleep research and complete the wearable validation series underpinning multimodal psychiatric biomarker studies using the Wearanize+ dataset.

## Data Availability Statement

Restrictions apply to the availability of these data. Data were obtained from the Wearanize+ project (Radboud University Medical Center, Donders Institute for Brain, Cognition and Behaviour, Nijmegen, The Netherlands) and are available at doi.org/10.34973/j6jf-9e62 with the permission of the Wearanize+ data custodians upon signing a Data Use Agreement.Analysis code used to produce the results reported in this manuscript is available on request from the corresponding author.

## IRB Statement

This study used a publicly available secondary dataset (Wearanize+ PlugNPlay, Radboud Data Repository). All data collection, participant recruitment, and ethical oversight were conducted by the original dataset creators. The original study was approved by the relevant institutional ethics committee and all participants provided written informed consent prior to data collection, as described in Sikder et al. (2026). Secondary analysis of this anonymised publicly available dataset under a signed Data Use Agreement did not require additional ethical approval from the authors’ institutional review board.

